# Clinical value of plasma ALZpath pTau217 immunoassay for assessing mild cognitive impairment

**DOI:** 10.1101/2024.01.21.24301570

**Authors:** Sylvain Lehmann, Susanna Schraen-Maschke, Jean-Sébastien Vidal, Constance Delaby, Luc Buée, Frédéric Blanc, Claire Paquet, Bernadette Allinquant, Stéphanie Bombois, Audrey Gabelle, Olivier Hanon, the BALTAZAR study group

**Author notes:** ***Corresponding author*** Pr Sylvain Lehmann, CHU and University of Montpellier, 80 av Fliche, 34295 Montpellier, France.

## Abstract

**Objectives:** We sought to compare two of the most promising plasma biomarkers for Alzheimer disease (AD): pTau181 and pTau217.

**Methods:** pTau181 and pTau217 were quantified using, Quanterix and ALZpath, SIMOA assays in the well characterized prospective multicentre BALTAZAR cohort of mild cognitive impairment (MCI) participants.

**Results:** Among MCI participants, 55% were Aβ+ and 29% developed dementia due to AD. pTau181 and pTau217 were higher in the Aβ+ population with fold-change of 1.5 and 2.7 respectively. MCI that converted to AD also had higher levels than non-converters, with hazard ratios of 1.38 (1.26-1.51) for pTau181 compared to 8.22 (5.45-12.39) for pTau217. The AUC for predicting Aβ+ was 0.783 (95%CI: 0.721-0.836; cut-point 2.75 pg/mL) for pTau181 and 0.914 (95%CI: 0.868-0.948; cut-point 0.44 pg/mL) for pTau217. The high predictive power of pTau217 was not improved by adding age, sex, and APOEε4 status, in a logistic model. Age, APOEε4 and renal dysfunction were associated with pTau levels, but the clinical performance of pTau217 was only marginally altered by these factors. Using a two cut-point approach, a 95% positive predictive value for Aβ+ corresponded to pTau217 > 0.8 pg/mL and a 95% negative predictive value at <0.23 pg/mL. At these two cut-points, the percentages of MCI conversion were 56.8% and 9.7% respectively, while the annual rates of decline in MMSE were −2.32 versus −0.65.

**Conclusions:** Plasma pTau217 and pTau181 both correlate with AD, but the fold-change in pTau217 make it better to diagnose cerebral amyloidosis, and predict cognitive decline and conversion to AD dementia.

## Introduction

Alzheimer disease (AD) is a problem that needs close monitoring for better management in an aging society where it is increasingly prevalent. AD likely follows a trajectory; amyloid build-up is thought to be the preclinical starting point in cognitively unimpaired people. These people start to have cognitive problems in this ‘prodromal’ stage. This mild cognitive impairment (MCI) is associated with gradual build-up of neuronal Tau tangles and conversion to dementia due to AD. There is an urgent need for the use in clinical practice of biomarkers for these early stages to better manage the disease.

Tau protein is at the heart of AD, and it exists in many post-translationally modified protein isotypes. Tau has many phosphorylation sites, many of which are in the proline-rich region, and some of these have been posited as useful biomarkers ^1^. Two of the moieties that have generated the most interest are threonines 181 and 217. Although pTau181 is a useful marker to predict amyloid status and conversion to dementia ^2^, many publications in the last three years are painting a picture whereby pTau217 is even more promising^3^. For example, in 2020 cerebrospinal fluid (CSF) pTau217 was already found to outperform pTau181 to detect AD ^45^. A likely crucial factor in this superiority was that the fold-change in CSF was greater for phosphorylation position 217 than for 181^6^.

The brain changes that occur in AD can currently be assessed by two methods: either by PET or by CSF analysis. It is thus essential to find blood biomarkers that mirror these more invasive/expensive/lengthy tests as a pre-screen. As mentioned above, many studies have also found plasma pTau217 highly specific in distinguishing AD from normal, e.g.^7–10^. Indeed, plasma pTau217 is as good as any CSF markers or as PET screening to discriminate AD from other diseases ^11^. pTau217 can also distinguish AD from other forms of dementia like FTLD ^10^. Even more importantly from a clinical perspective, plasma pTau217 can distinguish different stages of the AD trajectory.

Several studies have demonstrated a link between pTau217 and cerebral amyloidosis (Aβ+). pTau217 has the power to detect Aβ+ MCI ^12^ and Aβ+ can be detected with an AUC of 0.91 ^13^. For example, Doré and colleagues found that preclinical subjects that are Aβ+ had twice the level of pTau217, rising to 3.5X in cognitive impairment ^14^. The Hansson group have shown that pTau217 correlates with clinical deterioration, cognitive decline and brain atrophy and can detect the difference between cognitively unimpaired Aβ+ and Aβ-, making it a surrogate marker for preclinical and prodromal AD ^3 15–17^. In fact, plasma pTau217 is a predictor of poor cognitive trajectory ^8 18^ and conversion to AD ^19 20^. Plasma pTau217 builds up for two decades before the onset of symptoms ^21^ and plasma pTau217 and 231 build-up earlier than Aβ PET can detect changes ^22^.

Two different plasma pTau217 immunoassays: p-tau217Lilly (run on a Meso Scale Discovery (MSD) platform) and p-tau217Janssen (run on a Simoa plateform) were compared with similar results by Groot et al ^23^, and Janelidze et al ^12^. These assays were developed with proprietary antibodies. These results were further confirmed although with some doubts about their sensitivity to detect pTau217 all the time ^24 25^. More recently, novel pTau217 assays (University of Gothenburg [UGOT]) run on Simoa using commercially available tau12 and tau-441 antibodies gave very good results ^13^.

Not all the pTau217 assays used in these studies are available off-the-shelf to all investigators. Indeed, the ALZpath plasma pTau217 assay, which we evaluate below, is in fact the first scalable commercially available test. An article preprint has already suggested it is accurate in detecting AD pathology ^26^. In our results presented below this assay is comparable to other pTau217 assays in identifying cerebral amyloidosis, cognitive decline and conversion to AD dementia. Moreover, our study integrates the evaluation of comorbidities since the BALTAZAR cohort includes biomarkers designed to monitor metabolism, nutrition, diabetes and cardiovascular risk. Importantly, unlike for pTau181, the impact of comorbidities on performance seems to be limited probably in relation with the high fold change observed between normal and pathological groups. We therefore propose useful thresholds to confirm or rule out the presence of cerebral amyloidosis, information that can be used to stratify patients to select those who will benefit from the last line of anti-amyloid treatment.

## Materials and Methods

### Study population

This study included MCI participants of the BALTAZAR multicenter prospective cohort (ClinicalTrials.gov Identifier #NCT01315639)^27^. All participants had clinical, neuropsychological, brain MRI and biological assessments (see below). Right and left hippocampal volumes were obtained for each participant using virtual segmentation of the hippocampus. APOE was genotyped in a single centralized laboratory. MCI subjects were selected according to the Petersen criteria ^28^. Participants were assessed for conversion to dementia every six months for three years ^27^. The progression from MCI to dementia was defined by evaluation of the following parameters: (i) decline in cognitive function measured by mini-mental state examination (MMSE), (ii) disability in activities of daily living (ADL) and (iii) clinical dementia rating sum of boxes. Conversions from MCI to AD dementia were reviewed by an adjudication committee. Conversion to AD accounted for 95% of conversions to dementia, and was assessed on the basis of clinical, imaging and neuropsychological evaluation and follow-up. Participants were categorized, as amyloid-positive (Aβ+) or negative (Aβ-), based on their cerebrospinal fluid Aβ42/Aβ40 (ratio below 10% as measured with Euroimmun ELISA assays). Blood and CSF samples were taken on the same day, and to minimize pre-analytical and analytical problems, identical plasma collection tubes were used across centers. Plasma aliquots were stored at −80°C until testing.

### Plasma pTau Measurement

Plasma pTau level was determined, using the Quanterix method that is based on ultrasensitive Simoa technology ^29^, on an HD-X analytical platform. Plasma pTau181 was measured with a commercial Advantage V1 kit (#104111). This assay has a low limit of detection at 0.019 pg/mL and a low limit of quantification at 0.085 pg/mL. Quality controls, with low (QC 1 with mean concentration of 3.82 pg/mL) or high (QC 2 - 52.4 pg/mL) assigned pTau181 concentrations, are provided in the kits. Inter-assay coefficients of variation for QC 1 and QC 2 were 7% and 5%, respectively. Plasma pTau217 was detected using a novel immunoassay developed by ALZpath, utilizing a proprietary monoclonal pTau217-specific antibody. For this assay, the low limit of detection was 0.0052 pg/mL and the limit of quantification was 0.06 pg/mL. Intra- and inter-run precision were 11.4% and 14.6%, respectively.

### Biological Biomarker Measurements

Blood samples, taken at baseline, were used for determination of routine parameters in ISO15189-certified laboratories: fasting glycemia, triglycerides, cholesterol (total, high-density lipoproteins (HDL), low-density lipoproteins (LDL)), creatinine, prealbumin, albumin, total protein, C-reactive protein (CRP), hemoglobin, vitamin B12, thyroid stimulating hormone (TSH), folate, and red-cell folate ^27^. Estimated glomerular filtration rate (eGFR) based on creatinine, age, and sex was calculated using the CKD Epidemiology Collaboration (CKD-EPI) equation, revised in 2021 without inclusion of race^30^. High molecular weight (HMW) adiponectin was measured on stored samples using the LUMIPULSE G platform.

### Statistical Analyses

General characteristics were analyzed in the MCI sample overall and in converter and non-converter MCI subsets. Categorical variables were analyzed as percentage and counts (% (N)), continuous variables as mean and standard deviation (M (SD)) or median [25-75 percentile IQR] and comparisons were made by χ^2^ test, t-test, Mann-Whitney U test or analysis of variance (ANOVA, Kruskal-Wallis test). Cox proportional hazards regression models for conversion, with time to dementia as a dependent variable, were computed, with adjustment for age at blood draw, sex, and APOEε4 allele carrier status. We additionally plotted Kaplan-Meier curves for the different pTau tertiles and differences between tertiles were calculated by Log rank test. For all analyses, a two-sided α-level of 0.05 was used for significance testing. Receiving Operator Characteristic (ROC) curves, using conversion as a dependent variable, were also used. The corresponding areas under the curve (AUCs) were compared using the Delong method ^31^. For each comparison, the size of the different groups is indicated in the Tables. Missing data have not been imputed. All analyses were performed using MedCalc (20·118) and R (R Core Team (2019)) software.

## Results

### Baseline MCI participant characteristics

Here we present data from 473 MCI patients from the BALTAZAR cohort ^27^ (Table 1). Mean age at baseline was 77.7 [SD 5.5] years. 28.5% of the subjects (135/473) converted to AD dementia during the 3-year period ^32^. Subjects who converted to AD dementia (MCI converters) did not differ from non-converters regarding their age, sex distribution, body mass index (BMI), or educational levels (Table 1). 39.1% (184/470) of the MCI participants were APOE ε4 carriers. The average MMSE score at baseline was 26.4 [SD 2.5] and MCI converters had lower MMSE at baseline and a much higher MMSE decline per year, at −3.45 [SD 4.26] on average vs. −0.42 [SD 1.89] for the non-converter population. Hippocampal volume (R+L) (cm^3^) was also lower in converters than in non-converters. Hippocampal volume was not correlated to plasma pTau levels (Supplementary Figure 1). All these differences remained significant after adjustment for age, sex, APOE ε4 and the educational status. pTau217 levels were always lower than pTau181 levels; respective mean plasma levels in the MCI population were 0.49 [SD 0.34] vs. 3.18 [SD 1.49] pg/mL. The two sets of values were however correlated (Pearson correlation coefficient 0.73 [CI 95% 0.68-0.77], significance level p<0.0001).

**Table 1.** Patient characteristics.

|  | Total<br>n<br>MCI | Value<br>(Mean(sd)) | Aβ- | Value<br>(Mean(sd)) | Aβ+ | Value<br>(Mean(sd)) | Aβ- vs<br>Aβ+<br>(p) | Adjusted<br>age, sex,<br>APOE<br>ε4,<br>education | n MCI<br>Non<br>Converter | Value<br>(Mean(sd)) | n MCI<br>Converter | Value<br>(Mean(sd)) | Converter<br>vs. non<br>converter<br>(p) | Adjusted<br>age, sex,<br>APOE<br>ε4,<br>education |
| --- | --- | --- | --- | --- | --- | --- | --- | --- | --- | --- | --- | --- | --- | --- |
| Age (years) | 473 | 77.7 (5.5) | 97 | 76.6 (5.1) | 116 | 78 (5.9) | 0.0780 | / | 338 | 77.4 (5.4) | 135 | 78.4 (5.7) | 0.0918 | / |
| Men (%) | 473 | 38.7 | 97 | 45.4 | 116 | 36.2 | 0.1782 | / | 338 | 38.4 | 135 | 39.4 | 0.7135 | / |
| BMI (kg/m<sup>2</sup>) | 466 | 25 (3.8) | 97 | 25.4 (3.7) | 112 | 24.2 (3.6) | 0.0207 | 0.2558 | 334 | 25.1 (3.8) | 132 | 24.7 (3.7) | 0.3021 | 0.8348 |
| MMSE (/30) | 462 | 26.4 (2.5) | 94 | 27.1 (2) | 113 | 25.8 (2.5) | <.0001 | 0.0003 | 328 | 26.7 (2.5) | 134 | 25.6 (2.5) | <.0001 | 0.0003 |
| MMSE / year | 417 | -1.38 (3.18) | 89 | -1.2 (2.87) | 105 | -1.87<br>(4.24) | 0.1902 | 0.1061 | 285 | -0.42 (1.9) | 132 | -3.45 (4.26) | <.0001 | <.0001 |
| 1 or 2 APOE4 alleles (%) | 470 | 39.1 | 97 | 15.5 | 116 | 53.4 | <.0001 | / | 335 | 31.9 | 135 | 57.0 | <.0001 | / |
| Hippocampal volume (R+L) (cm<sup>3</sup>) | 383 | 4.55 (1.12) | 82 | 4.64 (1.23) | 98 | 4.52 (0.95) | 0.4677 | 0.9674 | 271 | 4.79 (1.06) | 112 | 3.99 (1.06) | <.0001 | <.0001 |
| Educational level (years) | 472 | 5.2 (1.6) | 97 | 5.3 (1.5) | 115 | 5.4 (1.6) | 0.0277 | / | 337 | 5.2 (1.6) | 135 | 5.2 (1.6) | 0.6877 | / |
| pTtau217 (pg/mL) | 473 | 0.49 (0.34) | 97 | 0.28 (0.19) | 116 | 0.75 (0.34) | <.0001 | <.0001 | 338 | 0.41 (0.29) | 135 | 0.69 (0.37) | <.0001 | <.0001 |
| pTtau181 (pg/mL) | 473 | 3.18 (1.49) | 97 | 2.60 (1.42) | 116 | 3.87 (1.38) | <.0001 | <.0001 | 338 | 2.93 (1.39) | 135 | 3.81 (1.54) | <.0001 | <.0001 |

Values in the number (n) of MCI participants for which different data types were available and comparison between non-converter and converters, with t-student or $\chi^2$ and linear regression adjusted for age, sex, and the presence of the APOE ε4 allele; numbers were used to describe categorical variables. mean ± standard deviation for continuous variables.
Abbreviations - MCI: mild cognitive impairment; APOE: apolipoprotein E; BMI, body mass index; MMSE, Mini–Mental State Examination; sd, standard deviation.

### Plasma pTau217 and pTau181 in A***β***- and A***β***+ Participants

In the subgroup of MCI with available CSF amyloid measurements, participants could be stratified as Aβ- or Aβ+ according to their CSF Aβ42/Ab40 ratio. Both plasma pTau217 and pTau181 levels were higher in Aβ+MCI than in Aβ- (n= 116, 0.75 [SD 0.34] vs. n= 97, 0.28 [SD 0.19] pg/mL for pTau217 and 3.87 [SD 1.38] vs. 2.6 [SD 1.42] pg/mL for pTau181) (Figure 1AB, Table 1). This was also the case for CSF pTau181 (Aβ- 51.9 [SD 16.0] vs. Aβ+ 79.1 [SD 32.3] pg/mL). CSF pTau181 correlated better with plasma pTau217 than with pTau181 (Supplementary Figure 2). However, fold change was much higher for plasma pTau217 than for plasma pTau181 (2.67 vs. 1.48) as well as for CSF pTau181 (2.67 vs. 1.52) (Supplementary Table 1). The AUC for Aβ+ detection was significantly higher for pTau217 (0.914 (95% CI: 0.868-0.948) than for pTau181 (0.783 (95% CI: 0.721-0.836) (Figure 1, Table 2). Optimal cut-points were determined, by Youden index, at 0.44 pg/mL and 2.75 pg/mL for pTau217 and pTau181, respectively. The AUCs increased non-significantly in a logistic regression model with age, sex, and APOEε4 status (Figure 1CD, Table 2). Conversely, the predictive power of age, sex, and APOEε4 status was significantly improved by adding pTau217, with the AUC rising from 0.750 (95% CI: 0.686-0,807) to 0.931 (95% CI: 0.889-0,961). Regarding blood biomarker comorbidities, folate and CRP concentrations were slightly lower in the Aβ+ population (Supplementary Table 2). However, Bonferroni adjustment linked to the multiple comparison of comorbidities did not reach significance (p>0.001).

**Figure 1.**
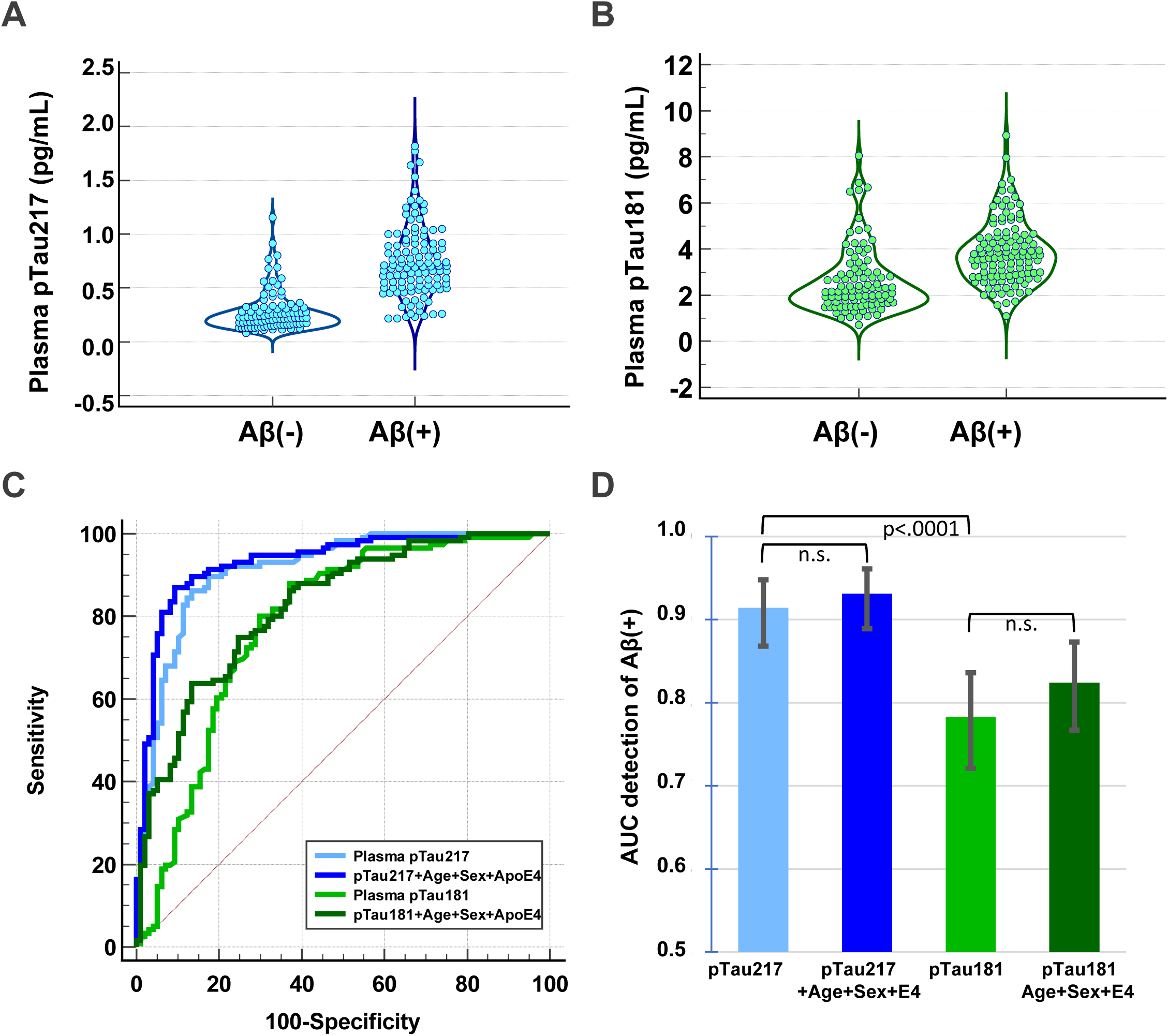
Plasma pTau217 and pTau181 in MCI according to amyloid status. Distribution of pTau217 (A) and pTau181 (B) in pg/mL are represented in the Aβ- and Aβ+ populations. (C) ROC curves for the same data. Both biomarkers were significantly different between these two populations and a logistic regression model combining pTau values with age, sex, and APOEε4 status gave slightly higher AUCs. (D) AUCs with 95% CIs.

**Table 2:**
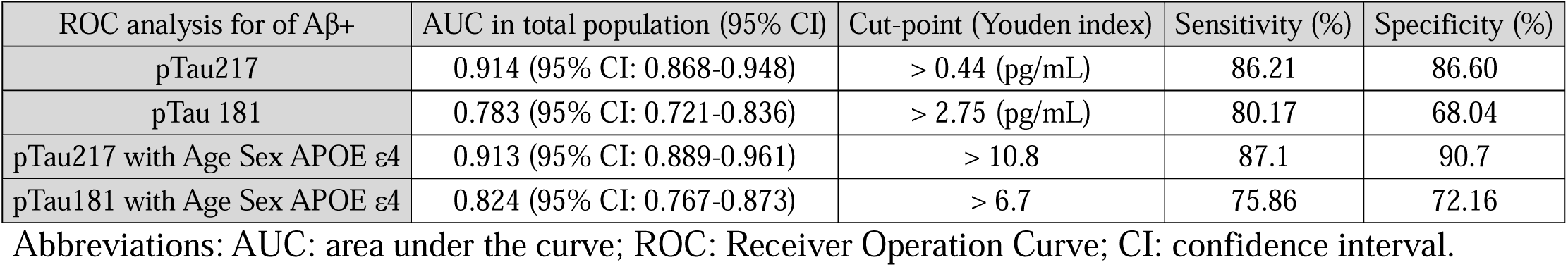
AUCs of ROC curves for Aβ+ detection.

### Plasma pTau217 and pTau181 predict cognitive decline and conversion to AD dementia

For MCI participants that converted to AD (n= 135) versus those that did not (n= 331), the respective values were 0.69 [SD 0.37] vs. 0.41 [SD 0.29] pg/mL for pTau217 and 3.81 [SD 1.53] vs. 2.93 [SD 1.39] pg/mL for pTau181, i.e. converters had 70% more pTau217 and only 30% more pTau218 (Table1, Supplementary Figure 3AB). The AUC for conversion to AD were significant, but they were lower than they were for the detection of cerebral amyloidosis: (0.746 (95% CI: 0.704-0.785) for pTau217 and (0.677 (95% CI: 0.633-0.719) for pTau181 (Supplementary Figure 3C). AUC for conversion, of CSF pTau181, was 0.712 (95% CI: 0.646-0.771), and CSF Aβ42/40: 0.733 (95% CI: 0.668-0.791). Participants who converted to AD were 78.3% Aβ+, whereas only 43.1% of non-converters were Aβ+. After adjustment for age, sex, and APOE ε4 status, in a Cox proportional hazard model, conversion to AD dementia, within three years, showed a significant risk for age, MMSE, APOE ε4, hippocampal volume, pTau181 and pTau217 (Table 3). pTau217 had a higher hazard ratio at 8.30 (5.46-12.61), compared to 1.38 (1.26-1.52) for pTau181. Importantly, none of the comorbidity biomarkers were independently associated with an increased risk of conversion (Supplementary Table 3). The relative risks of conversion to AD dementia, as predicted by high plasma pTau217 and pTau181, is illustrated by Kaplan-Meier curves of pTau tertiles (Figure 2AB). The hazard ratios (HRs) between the 1st and the 3rd tertile were 7.37 [95%CI: 4.86-11.16] and 3.83 [95%CI: 2.54 −5.79] for plasma pTau217 and pTau181, respectively. We also tracked changes in MMSE over 18 months (Figure 2CD) and found the steepest decline for the p217-high (third) tertile. The three p217 tertiles each predicted distinct cognitive decline trajectories. The differences were less significant for pTau181 with a smaller difference between low and medium and no further effect in the third pTau181 tertile.

**Figure 2.**
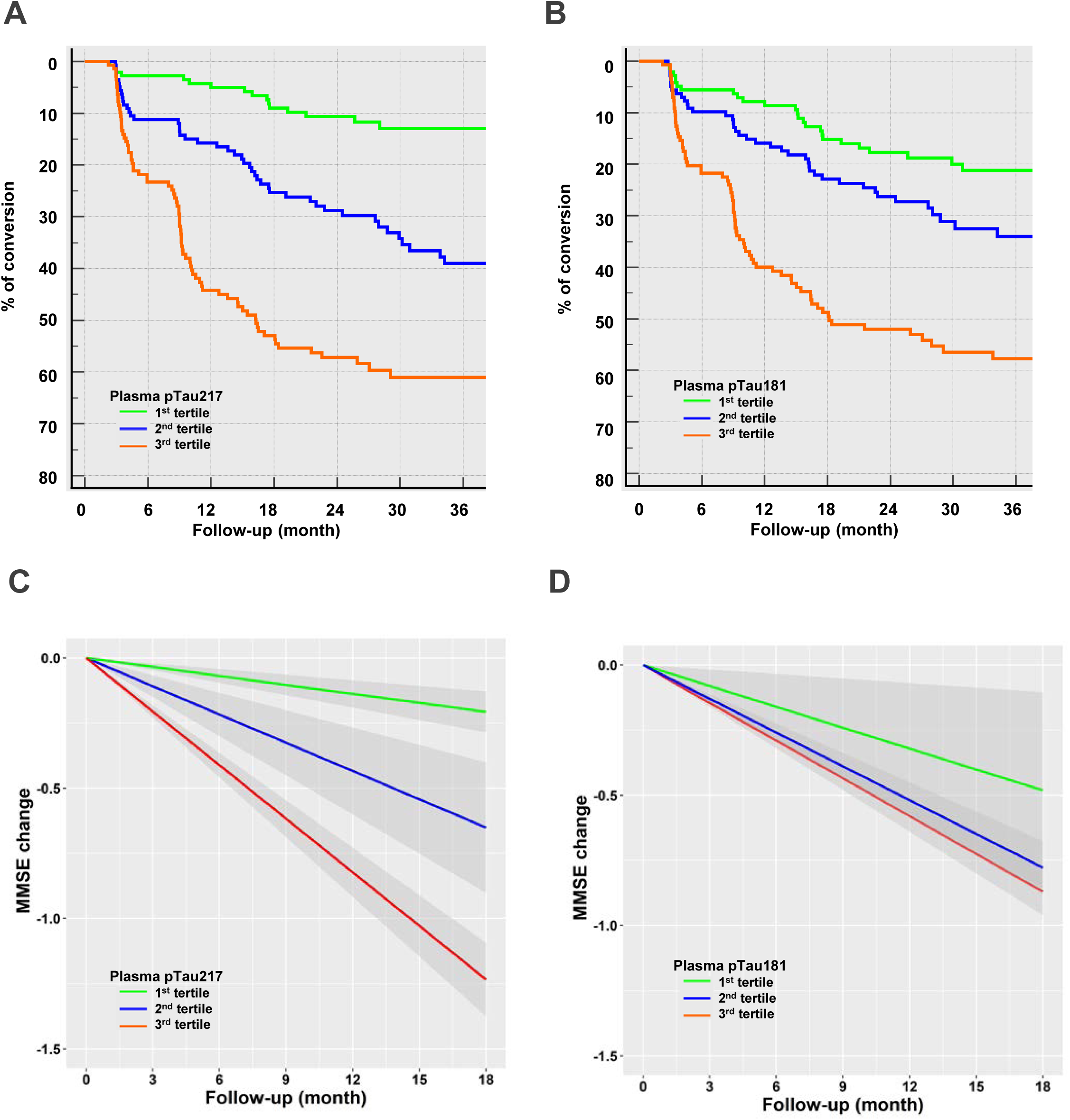
Conversion to AD dementia and MMSE evolution according to pTau181 or pTau217 tertiles. Plasma pTau217 and pTau181 measurements were separated into tertiles and conversion to AD dementia determined at six-month intervals over three years (Panels AB). A very significant overall difference was observed for both pTau217 and pTau181 (Logrank test (overall difference) 76.1 and 46.7 respectively, both P < 0.0001). Hazard ratio (HR) between 1st vs 3rd tertile was 7.37 (4.86 to 11.16) compared to just 3.83 (2.54 to 5.79 for plasma pTau217 and pTau181, respectively. The average slopes of MMSE decline per year in pTau terciles are plotted in panels CD. Grey shadows show the confidence interval. Lower lines show increasing tertile: first tertiles are green, second tertiles, blue, and third tertiles are orange.

**Table 3:** Risk factors associated with conversion to dementia during follow-up.

| Factors | n | HR Conversion (95%CI) | p | p adjusted (age, sex, APOE ε4) |
| --- | --- | --- | --- | --- |
| Age | 473 | 1.03 (1-1.07) | 0.0443 | / |
| Sex | 473 | 0.87 (0.62-1.23) | 0.4256 | / |
| BMI | 466 | 0.99 (0.95-1.04) | 0.7878 | 0.5496 |
| MMSE | 462 | 0.84 (0.79-0.89) | <0.0001 | <0.0001 |
| APOE ε4 | 470 | 2.34 (1.67-3.3) | <0.0001 | / |
| Hippocampal volume | 383 | 0.58 (0.5-0.67) | <0.0001 | <0.0001 |
| Educational level | 472 | 0.95 (0.85-1.05) | 0.3032 | 0.3014 |
| pTau217 | 473 | 8.30 (5.46-12.61) | <0.0001 | <0.0001 |
| pTau181 | 473 | 1,38 (1,26-1,52) | <0.0001 | <0.0001 |

Cox proportional hazard model of conversion to dementia in follow-up before and after adjustment for age, sex, educational level, and the APOE ε4 status. Abbreviations: HR, hazard ratio for conversion; CI: confidence interval; MCI, mild cognitive impairment; APOE: apolipoprotein E; BMI, body mass index; MMSE, Mini–Mental State Examination

### Association of plasma pTau217 and pTau181 levels with different biomarkers and cohort characteristics

The relationships, between plasma pTau concentrations and demographic or biological factors, collected at baseline in the BALTAZAR cohort, were studied using a linear regression approach. Plasma pTau217 and pTau181 were associated with BMI and APOE status (Figure 3A). The presence of APOE ε4 alleles was associated with significantly higher pTau values (t-test between APOE ε4 negative and positive population: p<0.0001). Levels of both pTau isoforms were also strongly related to renal function parameters: creatinine and eGFR (panel B). The only other biomarkers clearly associated with pTau levels were CRP for both isoforms and total protein for pTau181. The association between clinical chemistry analytes and plasma pTau levels was confirmed by calculation of Pearson correlations (Supplementary Table 4). To further assess the impact of renal function on pTau performance, participants were stratified, using eGFR values, between those having normal, slightly reduced or impaired renal function (Figure 3CD). Impaired renal function was associated with increased pTau values in both the Aβ- and Aβ+ MCI populations. However, renal function had a significant confounding impact on the performance of pTau181, with the optimal cut-point not separating the two populations well. On the other hand, renal parameters had little effect on plasma pTau217 performance, likely because this biomarker had a much higher fold change between the Aβ- and Aβ+ populations than pTau181.

**Figure 3.**
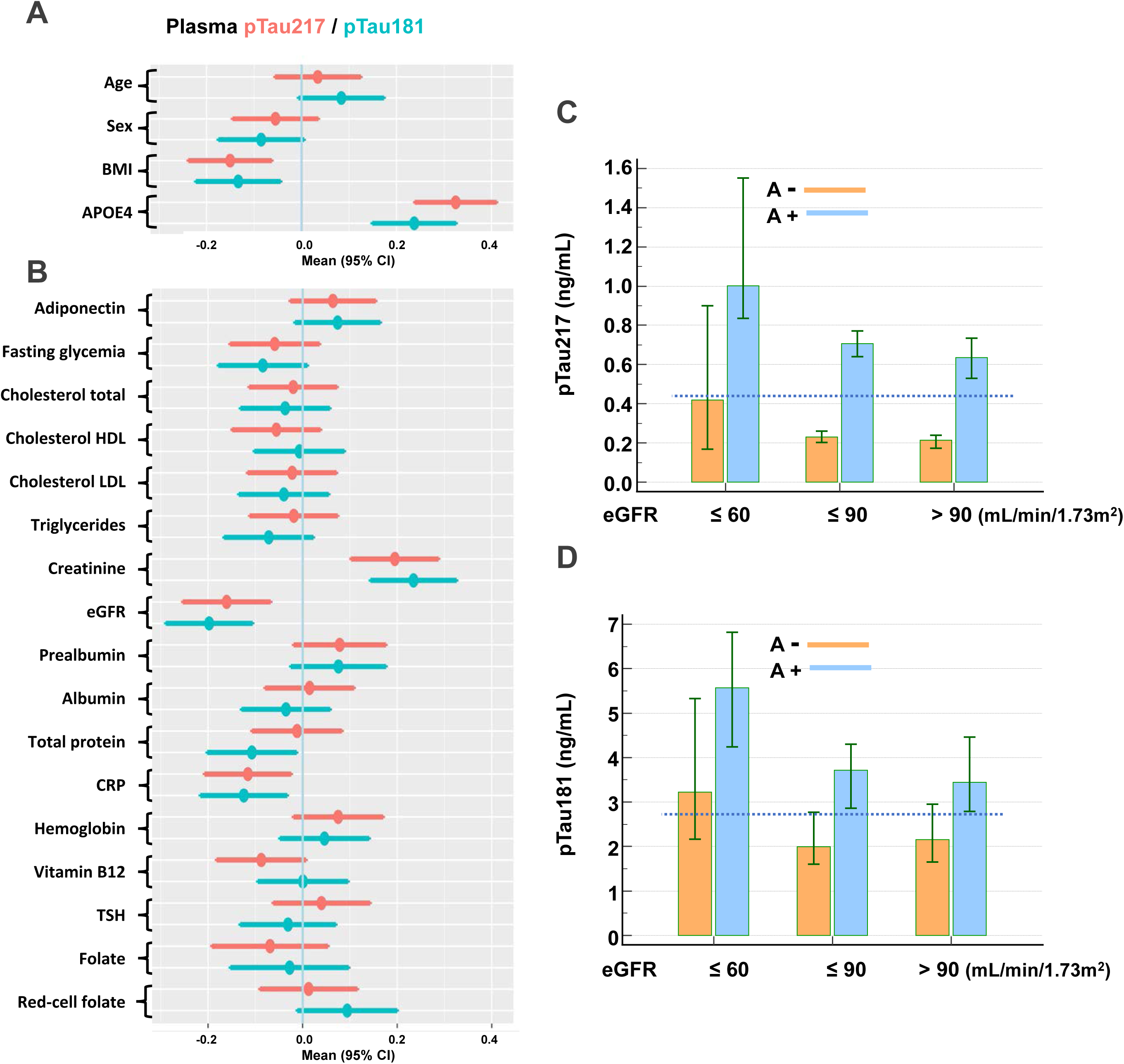
Association of plasma pTau217 and pTau181 levels with different biomarkers and cohort characteristics. Forest plots of associations between demographic (panel A) and comorbidity (panel B) biomarkers and plasma pTau217(red) or pTau181(blue), using linear regression of z-scores. Means and 95% confidence intervals (CIs) are provided. The concentrations of plasma pTau217 (panel C) or pTau181(panel D), in Aβ- (orange) and Aβ+ (blue) participants, are represented in participants stratified by their estimated glomerular filtration rate (eGFR), (eGFR<=60: impaired renal function; 60-90 mildly reduced renal function, >90 normal renal function). The value corresponding to the optimal cut-points for Aβ+ detection (Youden index) in all the population is represented by a dotted line. Note that the line separates the Aβ- and Aβ+ population for pTau217 only. Abbreviations: APOE, apolipoprotein E; BMI, body mass index; eGFR, estimated glomerular filtration rate; MMSE, Mini–Mental State Examination.

### Definition of pTau217 cut-point to detect cerebral amyloidosis

At the optimal threshold of 0.44 pg/mL, deduced from the ROC analysis, the positive predictive value (PPV) for Aβ+ detection was 88.5% and the negative predictive value (NPV) was 84.0% (Table 2). To achieve a PPV of 95%, plasma pTau217 had to be above 0.8 pg/mL, while to achieve an NPV of over 95%, pTau217 must be below 0.23 pg/mL. At these two cut-points, the percentages of MCI converting to AD dementia, over the three years, were 56.8% and 9.7% respectively, while the annual rates of decline in MMSE were −2.32 and −0.65 (Table 4).

**Table 4.** Characteristics of MCI participants at different pTau217 cutpoints.

| pTau217 cut-points (pg/mL) | % of MCI population | PPV of Aβ+ (%) | NPV of Aβ+ (%) | % AD dementia conversion | MMSE/year |
| --- | --- | --- | --- | --- | --- |
| > 0.44 | 53.1 | 88.5 (95% CI: 82.2-92.8) | 84.0 (95% CI: 76.8-89.3) | 46.3 | -2.07 |
| > 0.80 | 20.3 | 95.3 (95% CI: 83.6-98.8) | 55.9 (95% CI: 52.5-59.2) | 56.8 | -2.32 |
| < 0.23 | 24.4 | 70.8 (95% CI: 66.4-74.9) | 96.2 (95% CI: 86.2-99.0) | 9.7 | -0.65 |

Abbreviations: Positive and negative predictive values: PPV, NPV; MMSE, Mini–Mental State Examination

## Discussion

Here we examined the performance of plasma pTau181 and pTau217 in monitoring Alzheimer’s disease (AD) parameters, in the BALTAZAR cohort ^27^. This included data on 473 participants with mild cognitive impairment (MCI) over a three-year period, with regular assessments and biological fluid tests. Our main finding was that plasma pTau217 can accurately assess the presence of cerebral amyloidosis, with a confidence level above 95%. There was no relationship with hippocampal atrophy. Moreover, this biomarker predicts cognitive decline and the conversion of MCI to Alzheimer’s-type dementia. Plasma ALZpath pTau217 performs significantly better than the plasma pTau181 Advantage Simoa assay and overall, our results confirm previous observations of the superiority of pTau217 in other large cohorts ^3 18 25^. In our hands, plasma pTau217 matches CSF biomarkers for the prediction of conversion to AD and is even more effective than CSF pTau181 in identifying cerebral amyloidosis, consistent with recent work ^33^. This suggests that blood tests are comparable to or may even be more accurate than CSF tests.

However, before pTau217 assays can move to the clinic they will to have proven accuracy and they will also need to be made available for purchase and installation at a reasonable cost. Most published studies on pTau217 had one drawback; the tests were not available to standard clinical laboratories. These studies used in-house or proprietary assays that were not commercially available. With the growing interest in pTau217, this picture is set to evolve rapidly. Our data therefore represent a step forward in the deployment of clinical tests, which will eventually comply with the ISO15189 standard.

As well as clinical reproducibility, interpreting plasma pTau217 levels will require knowledge of the medical context, to be useful. An important factor for real-life use of assays in large populations is knowledge of the confounding factors that induce bias in the measurements. Indeed, it has been established that certain comorbidities, and in particular impaired renal function, can significantly modify the predictive value of plasma biomarkers. Other parameters such as age or BMI can also confound the value of AD prognostic biomarkers. For pTau181, we previously found impaired renal function likely undermines diagnostic performance ^34^. In the same cohort, we observed here that renal function and other potential confounding factors have a minimal effect on the performance of pTau217. This is likely due to the high fold-difference observed between normal and pathological situations. These results pave the way for wider, independent use of this marker. Note that none of the other factors we tested, such as age, sex, BMI, level of education or ApoE [4 genoytpe, either separately or together, significantly improve the independent predictive value of plasma pTau207, by more than an AUC of 0.02.

We are therefore in a situation where pTau217 alone can provide significant information for patient management, not only with regard to the presence of cerebral amyloidosis, which is important when selecting an anti-amyloid treatment and for diagnostic strategy ^35^, but also, for prognosis. Indeed, we demonstrate that high plasma pTau217 levels are associated with a high risk of conversion to AD dementia within three years. We can also see that cognitive evolution can be stratified with this marker, which is important information for the clinician.

For clinical use, it is also necessary to define one or more pathological cut-points, and recent papers have proposed for pTau217 different approaches depending on the medical need ^3 26 36 37^. A universally applicable plasma pTau217 cutpoint would therefore be useful for the management of patients presenting cognitive disorders. General practitioners would greatly appreciate a threshold ‘diagnosing’ cerebral amyloidosis with a 95% confidence level. In our cohort, the cut-point of > 0.44 pg/mL is a useful combination of sensitivity and specificity (>85%) and the cut-point of > 0.8 pg/mL gives a 95% positive predictive value for cerebral amyloidosis. Conversely, it is also very useful for patient management to be able to exclude the presence of cerebral amyloidosis with a high confidence (>95%). In our case, this low cut-point value is < 0.23 pg/mL. The intermediate zone of our study, or grey zone, between these two thresholds, represents 55% of the population. This percentage seems higher than in previous studies ^3^. The explanation certainly lies in our study population, which includes only MCI participants, and in the methods for detecting Aβ+ and pTau217.

The present study has some limitations. To increase the likelihood of conversion to AD we excluded participants with Lewy Body, Parkinson, frontotemporal or vascular MCI disorders. Therefore 77% of subjects had amnestic MCI and 28% of participants developed AD dementia. Amyloid status was available in only a part of the population, since the BALTAZAR study focused on conversion, and it was defined using CSF biomarkers rather than with PET amyloid. Conversion to AD was assessed using clinical, imaging and neuropsychological data, which represents a risk of error but avoids circular thinking about the use of biomarkers. The main strengths of the study lie in the large sample size of MCI participants that are well described, the controlled pre-analytical conditions, the use of a commercially available plasma pTau217 assays and the consideration of clinical chemistry analyte measurements realized at baseline.

## Conclusion

These data place us at the dawn of a major change in the management of Alzheimer’s Disease. This is linked to the clinical use of the plasma marker pTau217, whose performance, using commercially available assays is exceptional, both in terms of identifying cerebral amyloidosis and, as we have shown in this article, in predicting progression to Alzheimer’s dementia and accelerated cognitive decline. This information is essential for optimal patient management, including diagnostic strategy, prevention and access to disease modifying therapy.

## Statements

### Authors’ contributions

S.L., J.S.V. and O.H. take responsibility for the integrity of the data and the accuracy of the data analysis.

Concept and design: O.H., S.B., A.G., S S-M., S.L.

Acquisition, analysis, or interpretation of data: All authors.

Drafting of the manuscript: S.L.

Critical revision of the manuscript for important intellectual content: All authors.

Statistical analysis: S.L., J.S.V.

Obtained funding: O.H., S.B., A.G., S S-M., C.P., S.L.

All authors had full access to the data and contributed to revision and editing of the manuscript.

### Data availability

Data and informed consent form are available upon request after publication (APHP, Paris). Requests will be considered by each study investigator, based on the information provided by the requester, regarding the study and analysis plan. If the use is appropriate, a data sharing agreement will be put in place before distributing a fully de-identified version of the dataset, including the data dictionary used for analysis with individual participant data.

### Ethical appoval

All participants gave written informed consent to be part of the study. The BALTAZAR study was approved by the Paris ethics committee (CPP Ile de France IV Saint-Louis Hospital). Clinical trial registration: NCT01315639

### BALTAZAR study group

The BALTAZAR study group: Olivier Hanon [1], Frédéric Blanc [2], Yasmina Boudali [1], Audrey Gabelle [3], Jacques Touchon [3], Marie-Laure Seux [1], Hermine Lenoir [1], Catherine Bayle [1], StéphanieBombois [4], Christine Delmaire [4], Xavier Delbeuck [5], Florence Moulin [1], Emmanuelle Duron [6], Florence Latour [7], Matthieu Plichart [1], Sophie Pichierri [8], Galdric Orvoën [1], Evelyne Galbrun [9], Giovanni Castelnovo [10], Lisette Volpe-Gillot [11], Florien Labourée [1], Pascaline Cassagnaud [12], Claire Paquet [13], Françoise Lala [14], Bruno Vellas [14], Julien Dumurgier [13], Anne-Sophie Rigaud [1], Christine Perret-Guillaume [15], Eliana Alonso [16], Foucaud du Boisgueheneuc [17], Laurence Hugonot-Diener [1], Adeline Rollin-Sillaire [12], Olivier Martinaud [18], Clémence Boully [1], Yann Spivac [19], Agnès Devendeville [20], Joël Belmin [21], Philippe Robert [22], Thierry Dantoine [23], Laure Caillard [1], David Wallon [24], Didier Hannequin [18], Nathalie Sastre [14], Sophie Haffen [25], Anna Kearney-Schwartz [15], Jean-Luc Novella [26], Vincent Deramecourt [12], Valérie Chauvire [27], Gabiel Abitbol [1], Nathalie Schwald [19], Caroline Hommet [28], François Sellal [29], Marie-Ange Cariot [16], Mohamed Abdellaoui [30], Sarah Benisty [31], Salim Gherabli [1], Pierre Anthony [29], Frédéric Bloch [32], Nathalie Charasz [1], Sophie Chauvelier [1], Jean-Yves Gaubert [1], Guillaume Sacco [22], Olivier Guerin [22], Jacques Boddaert [33], Marc Paccalin [17], Marie-Anne Mackowiak [12], Marie-Thérèse Rabus [9], Valérie Gissot [34], Athanase Benetos [15], Candice Picard [20], Céline Guillemaud [35], Gilles Berrut [8], Claire Gervais [22], Jacques Hugon [13], Jean-Marc Michel [29], Jean-Philippe David [19], Marion Paulin [12], Pierre-JeanOusset [14], Pierre Vandel [36], Sylvie Pariel [21], Vincent Camus [37], Anne Chawakilian [1], Léna Kermanac’h [1], Anne-Cécile Troussiere [12], Cécile Adam [23], Diane Dupuy [20], Elena Paillaud [16], Hélène Briault [9], Isabelle Saulnier [38], Karl Mondon [37], Marie-Agnès Picat [23], Marie Laurent [16], Olivier Godefroy [20], RezkiDaheb [16], Stéphanie Libercier [29], Djamila Krabchi [1], Marie Chupin [39], JeanSébastien Vidal [1], Edouard Chaussade [1], Christiane Baret-Rose [40], Sylvain Lehmann [41], Bernadette Allinquant [40], Susanna Schraen-Maschke [4].

Affiliations

[1] EA 4468, APHP, Hospital Broca, Memory Resource and Research Centre of de Paris-Broca-Ile de France, Université Paris Cité, F-75013 Paris, France [2] CHRU de Strasbourg, Memory Resource and Research Centre of Strasbourg/Colmar, French National Centre for Scientific Research, ICube Laboratory and Fédération de Médecine Translationnelle de Strasbourg, Team Imagerie Multimodale Intégrative en Santé /Neurocrypto, Université de Strasbourg, F-67000 Strasbourg, France [3] Memory Research and Resources Center, Department of Neurology, Inserm INM NeuroPEPs Team, Université de Montpellier, F-34000 Montpellier, France [4] Inserm, CHU Lille, UMR-S-U1172, LiCEND, Lille Neuroscience & Cognition, LabEx DISTALZ, University of Lille, F-59000 Lille, France [5] Univ. Lille, Inserm U1171 Degenerative and Vascular Cognitive Disorders, F-59000 Lille, France. [6] Université Paris-Saclay, APHP, Hôpital Paul Brousse, département de gériatrie, Équipe MOODS, Inserm 1178, F-94800 Villejuif, France. [7] Centre Hospitalier de la Côte Basque, Department of Gerontology, F-64100 Bayonne, France. [8] Université de Nantes, EA 4334 Movement-Interactions-Performance, CHU Nantes, Memory Research Resource Center of Nantes, Department of clinical gerontology, F-44000 Nantes, France. [9] Sorbonne Université, APHP, Centre Hospitalier Dupuytren, Department of Gérontology 2, F-91210 Draveil, France. [10] CHU de Nimes, Hôpital Caremeau, Neurology Department, F-30029 Nimes, France. [11] Hôpital Léopold Bellan, Service de Neuro-Psycho-Gériatrie, Memory Clinic, F-75014 Paris, France. [12] Univ. Lille, CHU de Lille, Memory Resource and Research Centre of Lille, Department of Neurology, F-59000 Lille, France. [13] GHU APHP Nord Lariboisière Fernand Widal, Centre de Neurologie Cognitive, Université Paris Cité, F-75010 Paris, France [14] Université de Toulouse III, CHU La Grave-Casselardit, Memory Resource and Research Centre of Midi-Pyrénées, F-31300 Toulouse, France. [15] Université de Lorraine, CHRUdeNancy, Memory Resource and Research Centre of Lorraine, F-54500 Vandoeuvre-lès-Nancy, France. [16] Université de Paris, APHP, Hôpital européen Georges Pompidou, Service de Gériatrie, F-75015, Paris, France. [17] CHU de Poitiers, Memory Resource and Research Centre of Poitiers, F-86000 Poitiers, France. [18] CHU Charles Nicolle, Memory Resource and Research Centre of Haute Normandie, F-76000 Rouen, France. [19] APHP, Centre Hospitalier Émile-Roux, Department of Gérontology 1, F-94450 Limeil-Brévannes, France. [20] CHU d’Amiens-Picardie, Memory Resource and Research Centre of Amiens Picardie, F-80000 Amiens, France. [21] Sorbonne Université, APHP, Hôpitaux Universitaires Pitie-Salpêtrière-Charles-Foix, Service de Gériatrie Ambulatoire, F-75013 Paris, France. [22] Université Côte d’Azur, CHU de Nice, Memory Research Resource Center of Nice, CoBTek lab, F-06100 Nice, France. [23] CHU de Limoges, Memory Research Resource Center of Limoges, F-87000 Limoges, France. [24] Normandie Univ, UNIROUEN, Inserm U1245, CHU de Rouen, Department of Neurology and CNR-MAJ, Normandy Center for Genomic and Personalized Medicine, CIC-CRB1404, F-76000, Rouen, France. [25] CHU de Besançon, Memory Resource and Research Centre of Besançon Franche-Comté, F-25000 Besançon, France. [26] Université de Reims Champagne-Ardenne, EA 3797, CHU de Reims, Memory Resource and Research Centre of Champagne-Ardenne, F-51100 Reims, France. [27] CHU d’Angers, Memory Resource and Research Centre of Angers, F-49000 Angers, France. [28] CHRU de Tours, Memory Resource and Research Centre of Tours, F-37000 Tours, France. [29] Université de Strasbourg, CHRU de Strasbourg, Memory Resource and Research Centre of Strasbourg/Colmar, Inserm U-118, F-67000 Strasbourg, France. [30] Univ Paris Est Creteil, EA 4391 Excitabilité Nerveuse et Thérapeutique, CHU Henri Mondor, Department of Neurology, F-94000 Créteil, France. [31] Hôpital Fondation Rothschild, Department of Neurology, F-75019 Paris, France. [32] CHU d’Amiens-Picardie, Department of Gerontology, F-80000 Amiens, France. [33] Sorbonne Université, APHP, Hôpitaux Universitaires Pitie-Salpêtrière-Charles Foix, Memory Resource and Research Centre, Centre des Maladies Cognitives et Comportementales IM2A, Inserm UMR 8256, F-75013 Paris, France. [34] Université François-Rabelais de Tours, CHRU de Tours, MemoryResource andResearchCentre of Tours, Inserm CIC 1415, F-37000 Tours, France. [35] Sorbonne Université, APHP, Hôpitaux Universitaires Pitie-Salpêtrière-Charles Foix, Memory Resource and Research Centre, Centre des Maladies Cognitives et Comportementales IM2A, F-75013 Paris, France. [36] Université Bourgogne Franche-Comté, Laboratoire de Recherches Intégratives en Neurosciences et Psychologie Cognitive, CHU de Besançon, Memory Resource and Research Centre of Besançon Franche-Comté, F-25000 Besançon, France. [37] Université François-Rabelais de Tours, CHRU de Tours, UMR Inserm U1253, F-37000 Tours, France. [38] Université de Limoges, EA 6310 HAVAE, CHU de Limoges, Memory Research Resource Center of Limoges, F-87000 Limoges, France. [39] Université Paris-Saclay, Neurospin, CEA, CNRS, catineuroimaging.com, CATI Multicenter Neuroimaging Platform, F-91190 Gif-sur-Yvette, France. [40] Université de Paris, Institute of Psychiatric and Neurosciences, Inserm UMR-S 1266, F-75014 Paris, France. [41] Univ Montpellier, IRMB CHU de Montpellier, INM INSERM, Montpellier, France

## References

1. Wesseling H, Mair W, Kumar M, et al. Tau PTM Profiles Identify Patient Heterogeneity and Stages of Alzheimer’s Disease. Cell 2020;183(6):1699–713 e13. doi: 10.1016/j.cell.2020.10.029 [published Online First: 2020/11/15]

2. Lehmann S, Schraen-Maschke S, Vidal JS, et al. Plasma phosphorylated tau 181 predicts amyloid status and conversion to dementia stage dependent on renal function. J Neurol Neurosurg Psychiatry 2023;94(6):411–19. doi: 10.1136/jnnp-2022-330540 [published Online First: 2023/04/04]

3. Mattsson-Carlgren N, Salvado G, Ashton NJ, et al. Prediction of Longitudinal Cognitive Decline in Preclinical Alzheimer Disease Using Plasma Biomarkers. JAMA Neurol 2023;80(4):360–69. doi: 10.1001/jamaneurol.2022.5272 [published Online First: 2023/02/07]

4. Janelidze S, Stomrud E, Smith R, et al. Cerebrospinal fluid p-tau217 performs better than p-tau181 as a biomarker of Alzheimer’s disease. Nat Commun 2020;11(1):1683. doi: 10.1038/s41467-020-15436-0 [published Online First: 2020/04/05]

5. Barthelemy NR, Bateman RJ, Hirtz C, et al. Cerebrospinal fluid phospho-tau T217 outperforms T181 as a biomarker for the differential diagnosis of Alzheimer’s disease and PET amyloid-positive patient identification. Alzheimers Res Ther 2020;12(1):26. doi: 10.1186/s13195-020-00596-4 [published Online First: 2020/03/19]

6. Leuzy A, Janelidze S, Mattsson-Carlgren N, et al. Comparing the Clinical Utility and Diagnostic Performance of CSF P-Tau181, P-Tau217, and P-Tau231 Assays. Neurology 2021;97(17):e1681–e94. doi: 10.1212/WNL.0000000000012727 [published Online First: 2021/09/09]

7. Brickman AM, Manly JJ, Honig LS, et al. Plasma p-tau181, p-tau217, and other blood-based Alzheimer’s disease biomarkers in a multi-ethnic, community study. Alzheimers Dement 2021;17(8):1353–64. doi: 10.1002/alz.12301 [published Online First: 2021/02/14]

8. Pereira JB, Janelidze S, Stomrud E, et al. Plasma markers predict changes in amyloid, tau, atrophy and cognition in non-demented subjects. Brain 2021;144(9):2826–36. doi: 10.1093/brain/awab163 [published Online First: 2021/06/03]

9. Chen L, Niu X, Wang Y, et al. Plasma tau proteins for the diagnosis of mild cognitive impairment and Alzheimer’s disease: A systematic review and meta-analysis. Front Aging Neurosci 2022;14:942629. doi: 10.3389/fnagi.2022.942629 [published Online First: 2022/08/13]

10. Thijssen EH, La Joie R, Strom A, et al. Plasma phosphorylated tau 217 and phosphorylated tau 181 as biomarkers in Alzheimer’s disease and frontotemporal lobar degeneration: a retrospective diagnostic performance study. The Lancet Neurology 2021;20(9):739–52. doi: 10.1016/S1474-4422(21)00214-3 [published Online First: 2021/08/22]

11. Palmqvist S, Janelidze S, Quiroz YT, et al. Discriminative Accuracy of Plasma Phospho-tau217 for Alzheimer Disease vs Other Neurodegenerative Disorders. JAMA 2020;324(8):772–81. doi: 10.1001/jama.2020.12134 [published Online First: 2020/07/30]

12. Janelidze S, Bali D, Ashton NJ, et al. Head-to-head comparison of 10 plasma phospho-tau assays in prodromal Alzheimer’s disease. Brain 2023;146(4):1592–601. doi: 10.1093/brain/awac333 [published Online First: 2022/09/11]

13. Gonzalez-Ortiz F, Ferreira PCL, Gonzalez-Escalante A, et al. A novel ultrasensitive assay for plasma p-tau217: Performance in individuals with subjective cognitive decline and early Alzheimer’s disease. Alzheimers Dement 2023 doi: 10.1002/alz.13525 [published Online First: 2023/11/17]

14. Dore V, Doecke JD, Saad ZS, et al. Plasma p217+tau versus NAV4694 amyloid and MK6240 tau PET across the Alzheimer’s continuum. Alzheimers Dement (Amst) 2022;14(1):e12307. doi: 10.1002/dad2.12307 [published Online First: 2022/04/14]

15. Ashton NJ, Janelidze S, Mattsson-Carlgren N, et al. Differential roles of Abeta42/40, p-tau231 and p-tau217 for Alzheimer’s trial selection and disease monitoring. Nat Med 2022;28(12):2555-62. doi: 10.1038/s41591-022-02074-w [published Online First: 2022/12/02]

16. Mattsson-Carlgren N, Janelidze S, Palmqvist S, et al. Longitudinal plasma p-tau217 is increased in early stages of Alzheimer’s disease. Brain 2020;143(11):3234–41. doi: 10.1093/brain/awaa286 [published Online First: 2020/10/18]

17. Janelidze S, Berron D, Smith R, et al. Associations of Plasma Phospho-Tau217 Levels With Tau Positron Emission Tomography in Early Alzheimer Disease. JAMA Neurol 2021;78(2):149–56. doi: 10.1001/jamaneurol.2020.4201 [published Online First: 2020/11/10]

18. Jonaitis EM, Janelidze S, Cody KA, et al. Plasma phosphorylated tau 217 in preclinical Alzheimer’s disease. Brain Commun 2023;5(2):fcad057. doi: 10.1093/braincomms/fcad057 [published Online First: 2023/04/05]

19. Palmqvist S, Tideman P, Cullen N, et al. Prediction of future Alzheimer’s disease dementia using plasma phospho-tau combined with other accessible measures. Nature medicine 2021;27(6):1034–42. doi: 10.1038/s41591-021-01348-z [published Online First: 2021/05/26]

20. Groot C, Cicognola C, Bali D, et al. Diagnostic and prognostic performance to detect Alzheimer’s disease and clinical progression of a novel assay for plasma p-tau217. Alzheimers Res Ther 2022;14(1):67. doi: 10.1186/s13195-022-01005-8 [published Online First: 2022/05/16]

21. Barthelemy NR, Li Y, Joseph-Mathurin N, et al. A soluble phosphorylated tau signature links tau, amyloid and the evolution of stages of dominantly inherited Alzheimer’s disease. Nature medicine 2020;26(3):398–407. doi: 10.1038/s41591-020-0781-z [published Online First: 2020/03/13]

22. Mila-Aloma M, Ashton NJ, Shekari M, et al. Plasma p-tau231 and p-tau217 as state markers of amyloid-beta pathology in preclinical Alzheimer’s disease. Nat Med 2022;28(9):1797–801. doi: 10.1038/s41591-022-01925-w [published Online First: 20220811]

23. Groot C, Smith R, Stomrud E, et al. Phospho-tau with subthreshold tau-PET predicts increased tau accumulation rates in amyloid-positive individuals. Brain 2023;146(4):1580–91. doi: 10.1093/brain/awac329 [published Online First: 2022/09/10]

24. Bayoumy S, Verberk IMW, den Dulk B, et al. Clinical and analytical comparison of six Simoa assays for plasma P-tau isoforms P-tau181, P-tau217, and P-tau231. Alzheimer’s research & therapy 2021;13(1):198. doi: 10.1186/s13195-021-00939-9 [published Online First: 2021/12/06]

25. Palmqvist S, Stomrud E, Cullen N, et al. An accurate fully automated panel of plasma biomarkers for Alzheimer’s disease. Alzheimers Dement 2023;19(4):1204–15. doi: 10.1002/alz.12751 [published Online First: 2022/08/12]

26. Ashton NJ, Brum WS, Molfetta GD, et al. Diagnostic accuracy of the plasma ALZpath pTau217 immunoassay to identify Alzheimer’s disease pathology. medRxiv 2023:2023.07.11.23292493. doi: 10.1101/2023.07.11.23292493

27. Hanon O, Vidal JS, Lehmann S, et al. Plasma amyloid levels within the Alzheimer’s process and correlations with central biomarkers. Alzheimers Dement 2018;14(7):858–68. doi: 10.1016/j.jalz.2018.01.004 [published Online First: 2018/02/20]

28. Petersen RC, Smith GE, Waring SC, et al. Mild cognitive impairment: clinical characterization and outcome. Archives of neurology 1999;56(3):303–8. [published Online First: 1999/04/06]

29. Rissin DM, Walt DR. Digital concentration readout of single enzyme molecules using femtoliter arrays and Poisson statistics. Nano Lett 2006;6(3):520–3. doi: 10.1021/nl060227d [published Online First: 2006/03/09]

30. Inker LA, Eneanya ND, Coresh J, et al. New Creatinine- and Cystatin C-Based Equations to Estimate GFR without Race. N Engl J Med 2021;385(19):1737–49. doi: 10.1056/NEJMoa2102953 [published Online First: 2021/09/24]

31. DeLong ER, DeLong DM, Clarke-Pearson DL. Comparing the areas under two or more correlated receiver operating characteristic curves: a nonparametric approach. Biometrics 1988;44(3):837–45. [published Online First: 1988/09/01]

32. Hanon O, Vidal JS, Lehmann S, et al. Plasma amyloid beta predicts conversion to dementia in subjects with mild cognitive impairment: The BALTAZAR study. Alzheimers Dement 2022 doi: 10.1002/alz.12613 [published Online First: 2022/02/22]

33. Barthelemy NR, Salvado G, Schindler S, et al. Highly Accurate Blood Test for Alzheimer’s Disease Comparable or Superior to Clinical CSF Tests. Nat Med 2024 doi: 10.1038/s41591-024-02869-z [published Online First: 20240221]

34. Lehmann SS-M, S. JS Vidal, C Delaby, F Blanc, C Paquet, B Allinquant, S Bombois, A Gabelle, O Hanon. Plasma phosphorylated tau 181 predicts amyloid status and conversion to dementia stage dependent on renal function. J Neurol Neurosurg Psychiatry 2022;In press

35. Delaby C, Alcolea D, Hirtz C, et al. Blood amyloid and tau biomarkers as predictors of cerebrospinal fluid profiles. J Neural Transm (Vienna) 2022 doi: 10.1007/s00702-022-02474-9 [published Online First: 2022/02/17]

36. Brum WS, Cullen NC, Janelidze S, et al. A two-step workflow based on plasma p-tau217 to screen for amyloid beta positivity with further confirmatory testing only in uncertain cases. Nat Aging 2023;3(9):1079–90. doi: 10.1038/s43587-023-00471-5 [published Online First: 2023/09/01]

37. Mattsson-Carlgren N, Collij LE, Stomrud E, et al. Plasma Biomarker Strategy for Selecting Patients With Alzheimer Disease for Antiamyloid Immunotherapies. JAMA Neurol 2023 doi: 10.1001/jamaneurol.2023.4596 [published Online First: 20231204]

