## Supplementary material for "Clinical value of plasma ALZpath pTau217 immunoassay for assessing mild cognitive impairment"

#### Supplementary data

**Page 2: Supplementary Table 1:** Distribution of plasma pTau217, plasma pTau181 and CSF pTau181 in Aβ- and Aβ+ population

**Page 3: Supplementary Table 2.** Biological values in MCI population.

**Page 4: Supplementary Table 3:** Risk factors associated with conversion to dementia during follow up

**Page 5: Supplementary Table 4:** Correlation between plasma pTau181, pTau217 and clinical chemistry analytes

**Page 5: Supplementary Figure 1:** Correlation between plasma pTau217, plasma pTau181 and hippocampal volume.

**Page 6: Supplementary Figure 2:** Correlation between plasma pTau217 and pTau181 and CSF pTau181 in MCI according to amyloid status.

**Page 7: Supplementary Figure 3:** Distribution and ROC curves of plasma pTau217 and pTau181 in MCI according to conversion to AD.

#### Supplementary Table 1: Distribution of plasma pTau217, plasma pTau181 and CSF pTau181 in Aβ- and Aβ+ population

| Factors | n Aβ- | Value (Mean(sd)) | n Aβ+ | Value (Mean(sd)) | Aβ- vs Aβ+ (p) | Fold change |
|---|---|---|---|---|---|---|
| Plasma pTau217 (pg/mL) | 97 | 0.28 (0.19) | 116 | 0.75 (0.34) | <.0001 | 2.67 |
| Plasma pTau 181 (pg/mL) | 97 | 2.60 (1.42) | 116 | 3.87 (1.38) | <.0001 | 1.48 |
| CSF pTau181 (pg/mL) | 97 | 51.9 (16.0) | 116 | 79.1 (32.3) | <.0001 | 1.52 |

Values as mean ± standard deviation in the number (n) of MCI participants in Aβ- and Aβ+ population with t-student comparison test

### Supplementary Table 2. Biological values in MCI population.

| | Total n MCI | Value (Mean(sd)) | Aβ- | Value (Mean(sd)) | Aβ+ | Value (Mean(sd)) | Aβ- vs Aβ+ (p) | Adjusted age, sex, APOE ε4, education | n MCI Non Converter | Value (Mean(sd)) | n MCI Converter | Value (Mean(sd)) | Converter vs non converter (p) | Adjusted age, sex, APOE ε4, education |
|---|---|---|---|---|---|---|---|---|---|---|---|---|---|---|
| Adiponectin (ug/mL) | 472 | 8.86 (6.22) | 97 | 7.05 (4.25) | 116 | 8.59 (5.92) | 0.0279 | 0.1331 | 337 | 8.94 (6.49) | 135 | 8.64 (5.49) | 0.6037 | 0.3337 |
| Fasting glycemia (mmol/L) | 439 | 5.37 (1.19) | 90 | 5.42 (1.15) | 114 | 5.42 (1.16) | 0.9977 | 0.6896 | 311 | 5.33 (1.2) | 128 | 5.45 (1.17) | 0.3300 | 0.2035 |
| Triglycerides (mmol/L) | 421 | 5.5 (1.16) | 87 | 5.54 (1.26) | 107 | 5.5 (1.12) | 0.8158 | 0.2670 | 297 | 1.22 (0.6) | 124 | 1.22 (0.55) | 0.9890 | 0.8715 |
| Cholesterol total (mmol/L) | 419 | 1.74 (0.52) | 87 | 1.76 (0.49) | 108 | 1.71 (0.51) | 0.5282 | 0.7774 | 297 | 5.46 (1.2) | 124 | 5.57 (1.06) | 0.3540 | 0.4530 |
| Cholesterol HDL (mmol/L) | 417 | 3.2 (1) | 87 | 3.25 (1.07) | 107 | 3.25 (0.92) | 0.9933 | 0.1931 | 296 | 1.74 (0.53) | 123 | 1.73 (0.48) | 0.8324 | 0.6706 |
| Cholesterol LDL (mmol/L) | 421 | 1.22 (0.59) | 87 | 1.18 (0.48) | 108 | 1.15 (0.69) | 0.7116 | 0.6172 | 295 | 3.17 (1.02) | 122 | 3.28 (0.96) | 0.2794 | 0.4081 |
| Creatinine (µmol/L) | 443 | 78.21 (20.97) | 92 | 80.15 (24.88) | 113 | 78.31 (20.02) | 0.5657 | 0.7440 | 316 | 78.36 (20.47) | 127 | 77.84 (22.25) | 0.8216 | 0.4137 |
| eGFR (mL/min/1.73m2) | 444 | 76.99 (14.7) | 92 | 77.51 (15.55) | 114 | 76.69 (15.34) | 0.7061 | 0.5876 | 317 | 76.81 (14.94) | 127 | 77.44 (14.14) | 0.6766 | 0.4420 |
| Prealbumin (mg/dl) | 385 | 0.279 (0.061) | 84 | 0.285 (0.077) | 99 | 0.282 (0.05) | 0.7025 | 0.5216 | 268 | 0.28 (0.06) | 117 | 0.28 (0.06) | 0.4165 | 0.3961 |
| Albumin (g/L) | 442 | 40.32 (3.9) | 93 | 39.89 (3.59) | 110 | 40.19 (4.9) | 0.6183 | 0.3942 | 318 | 40.52 (3.53) | 124 | 39.81 (4.68) | 0.1279 | 0.1349 |
| Total protein (g/L) | 430 | 70.46 (6.67) | 90 | 70.69 (5.69) | 111 | 70.31 (4.1) | 0.5889 | 0.7118 | 305 | 70.33 (7.41) | 125 | 70.75 (4.38) | 0.4715 | 0.4367 |
| CRP (mg/L) | 438 | 3.21 (5.39) | 90 | 4.76 (8.71) | 111 | 2.28 (2.7) | 0.0107 | 0.0120 | 314 | 3.05 (4.59) | 124 | 3.63 (7.01) | 0.3959 | 0.1054 |
| Hemoglobin (g/dL) | 439 | 13.8 (1.15) | 91 | 13.76 (1.21) | 112 | 13.84 (0.95) | 0.5831 | 0.2239 | 312 | 13.75 (1.19) | 127 | 13.92 (1.04) | 0.1594 | 0.2466 |
| Vitamin B12 (pmol/L) | 408 | 310.9 (131.7) | 86 | 322.4 (156.2) | 101 | 307.5 (108.3) | 0.4559 | 0.2743 | 291 | 315.27 (137.18) | 101 | 300 (116.79) | 0.2576 | 0.4200 |
| TSH (mU/L) | 391 | 2.31 (2.69) | 83 | 2.29 (2.73) | 101 | 2.39 (3) | 0.8137 | 0.6952 | 274 | 2.25 (2.22) | 101 | 2.43 (3.57) | 0.6139 | 0.6643 |
| Folate (nmol/L) | 216 | 29.45 (47.32) | 32 | 40.44 (71.58) | 42 | 21.69 (24.3) | 0.1640 | 0.0238 | 156 | 30.03 (44.11) | 42 | 27.97 (55.17) | 0.7963 | 0.9238 |
| Red-cell folate (nmol/L) | 343 | 1239 (726) | 78 | 1404 (1015) | 88 | 1328 (656) | 0.5722 | 0.9222 | 240 | 1225 (737) | 88 | 1271 (701) | 0.5796 | 0.3599 |

Values in the number (n) of MCI participants for which different data types were available and comparison between non-converter and converters, with t-student or $\chi 2$ and linear regression adjusted for age, sex, and the presence of the APOE ε4 allele; numbers were used to describe categorical variables. mean ± standard deviation for continuous variables. After Bonferroni adjustment, the significant P value is set at 0.003. Abbreviations: MCI: mild cognitive impairment; eGFR, estimated glomerular filtration rate; sd, standard deviation.

#### Supplementary Table 3: Risk factors associated with conversion to dementia during follow up

| Factors | n | HR Conversion (95%CI) | p | p adjusted (age, sex, ApoE) |
|---|---|---|---|---|
| Adiponectin | 472 | 0.96 (1.02-1.02) | 0.5870 | 0.2982 |
| Fasting glycemia | 439 | 0.97 (1.27-1.27) | 0.1208 | 0.0540 |
| Triglycerides | 421 | 0.88 (1.64-1.64) | 0.2389 | 0.1680 |
| Cholesterol total | 421 | 0.92 (1.24-1.24) | 0.3886 | 0.1671 |
| Cholesterol HDL | 419 | 0.59 (1.17-1.17) | 0.2824 | 0.8413 |
| Cholesterol LDL | 417 | 0.95 (1.34-1.34) | 0.1835 | 0.1404 |
| Creatinine | 443 | 0.99 (1.01-1.01) | 0.7844 | 0.5356 |
| eGFR | 444 | 0.99 (1.01-1.01) | 0.9811 | 0.7385 |
| Prealbumin | 385 | 0.01 (5.35-5.35) | 0.3690 | 0.4187 |
| Albumin | 442 | 0.90 (0.99-0.99) | 0.0193 | 0.0272 |
| Total protein | 430 | 0.98 (1.04-1.04) | 0.7511 | 0.6173 |
| CRP | 438 | 0.99 (1.05-1.05) | 0.2197 | 0.0452 |
| Hemoglobin | 439 | 0.97 (1.31-1.31) | 0.1140 | 0.1336 |
| Vitamin B12 | 408 | 1.00 (1.00-1.00) | 0.2269 | 0.4278 |
| TSH | 391 | 0.97 (1.10-1.10) | 0.3234 | 0.637 |
| Folate | 216 | 0.99 (1.00-1.00) | 0.4958 | 0.8443 |
| Red-cell folate | 343 | 1.00 (1.00-1.00) | 0.9336 | 0.5171 |

Cox proportional hazard model of conversion to dementia in follow-up before and after adjustment for age, sex, educational level, and the APOE ε4 status.  Abbreviations: HR, hazard ratio for conversion; CI: confidence interval.

#### Supplementary Table 4: Correlation between plasma pTau181, pTau217 and clinical chemistry analytes

| | pTau181 | | | pTau217 | | |
|---|---|---|---|---|---|---|
| | **n** | **Correlation coefficient** | **Significance Level P** | **n** | **Correlation coefficient** | **Significance Level P** |
| **Adiponectin** | 472 | 0.074 | 0.1089 | 472 | 0.064 | 0.164 |
| **Fasting glycemia** | 439 | -0.084 | 0.0795 | 439 | -0.058 | 0.2219 |
| **Triglycerides** | 421 | -0.073 | 0.1358 | 421 | -0.02 | 0.6892 |
| **Cholesterol total** | 421 | -0.037 | 0.4432 | 421 | -0.021 | 0.6712 |
| **Cholesterol HDL** | 419 | -0.007 | 0.8866 | 419 | -0.057 | 0.242 |
| **Cholesterol LDL** | 417 | -0.04 | 0.4133 | 417 | -0.023 | 0.6392 |
| **Creatinine** | 444 | 0.236 | **<0.0001** | 444 | 0.188 | **0.0001** |
| **eGFR** | 444 | -0.24 | **<0.0001** | 444 | -0.187 | **0.0001** |
| **Prealbumin** | 385 | 0.075 | 0.1408 | 385 | 0.08 | 0.1163 |
| **Albumin** | 442 | -0.035 | 0.4587 | 442 | 0.015 | 0.7591 |
| **Total protein** | 438 | -0.126 | **0.0082** | 438 | -0.118 | **0.0134** |
| **CRP** | 430 | -0.107 | **0.0262** | 430 | -0.012 | 0.8069 |
| **Hemoglobin** | 439 | 0.046 | 0.3365 | 439 | 0.074 | 0.1203 |
| **Vitamin B12** | 408 | 0.001 | 0.9899 | 408 | -0.09 | 0.0698 |
| **TSH** | 391 | -0.031 | 0.5475 | 391 | 0.039 | 0.4444 |
| **Folate** | 216 | -0.03 | 0.6651 | 216 | -0.075 | 0.2693 |
| **Red-cell folate** | 343 | 0.095 | 0.0805 | 343 | 0.013 | 0.8078 |

Pearson's correlation coefficient in MCI particpants (n) between clinical chemistry analytes and plasma pTau levels.

**Supplementary Figure 1: Correlation between plasma pTau217, plasma pTau181 and hippocampal volume.**

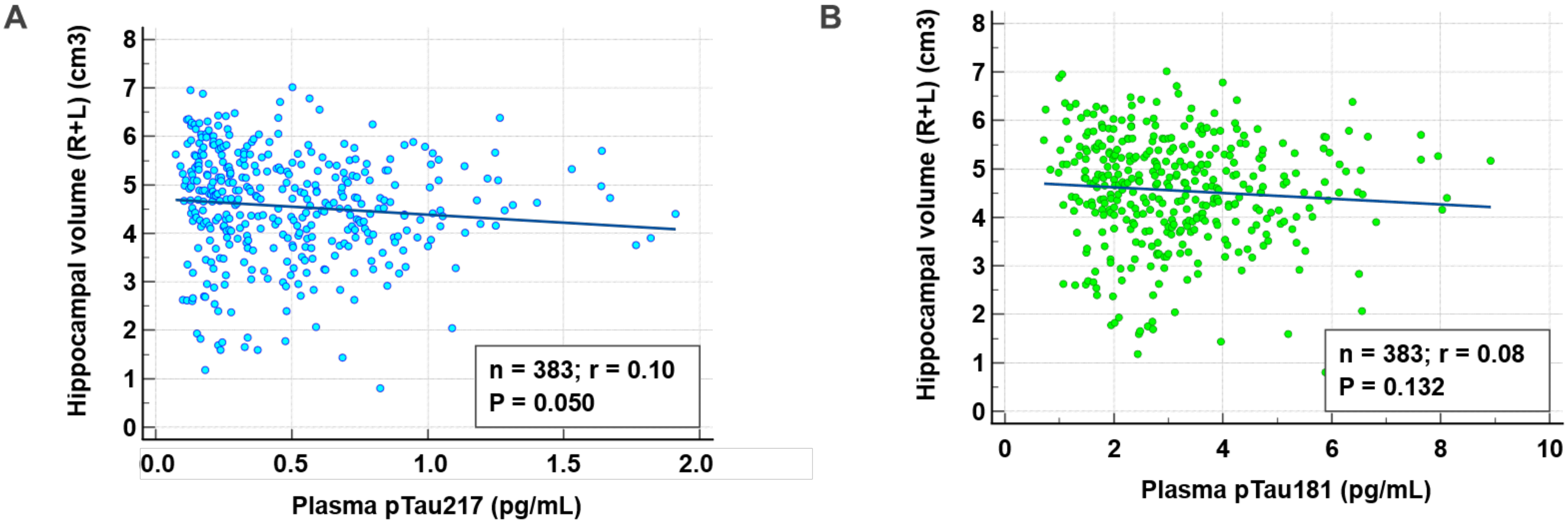


**Legend:**

Values of pTau217 (A) and pTau181 (B) in pg/mL were correlated with Hippocampal volume (R+L) (cm3). Pearson's correlation coefficient 'r' are indicated and they did not reach significance.

#### Supplementary Figure 2: Correlation between plasma pTau217 and pTau181 and CSF pTau181.

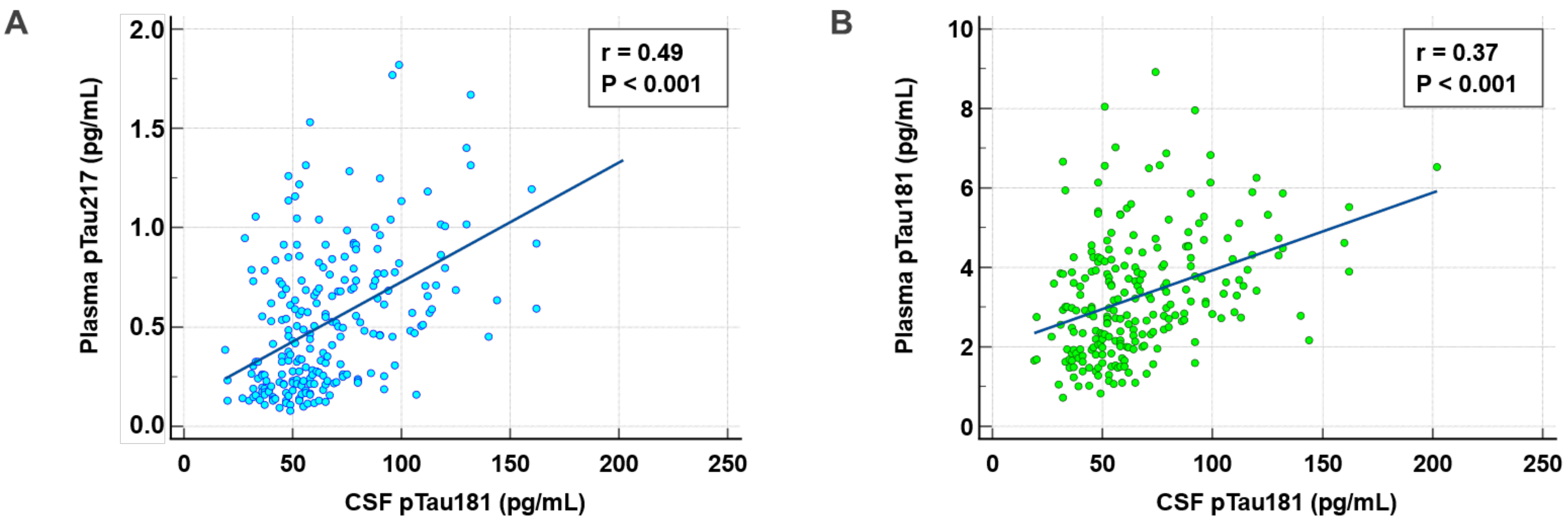


**Legend:**

Correlation between plasma pTau217 (Panel A) and pTau181 (Panel B) and CSF pTau181 in MCI according to amyloid status. Pearson's correlation coefficients 'r' were 0.49 (95% Confidence interval 0.38 to 0.58) and 0.37 (95% Confidence interval 0.26 to 0.48) for plasma pTau217 vs. CSF pTau181, and plasma pTau181 vs. CSF pTau181, respectively.

**Supplementary Figure 3: Distribution of plasma pTau217 and pTau181 and ROC curves of plasma pTau217, plasma pTau181 and CSF pTau181 and CSF Aβ42/40 in MCI according to conversion to AD.**

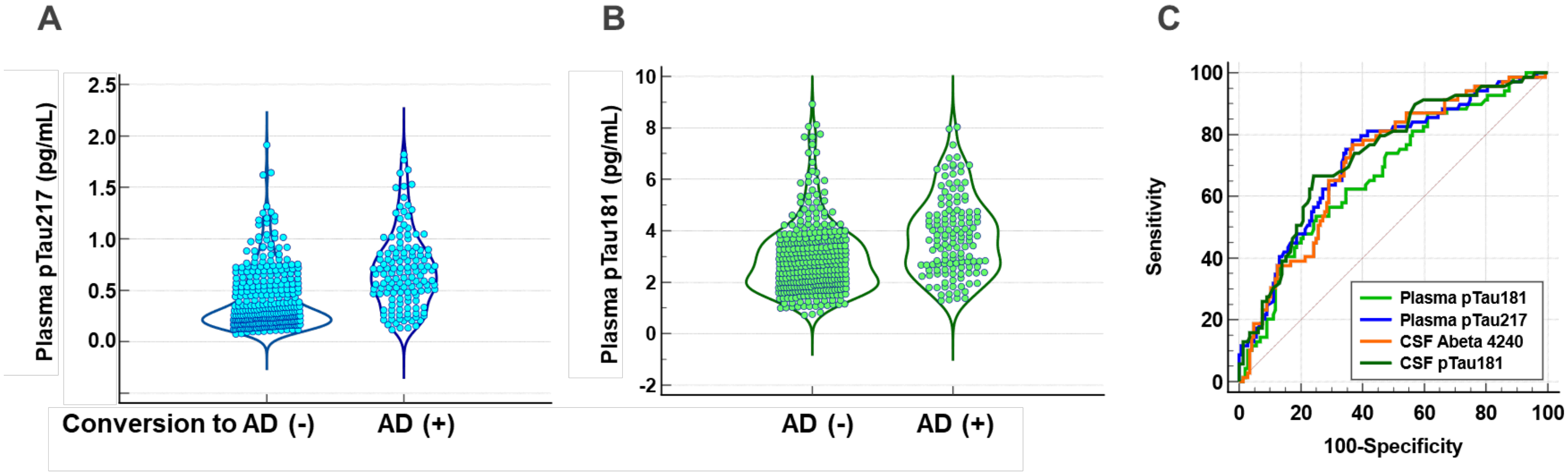


**Legend:**

Values of pTau217 (A) and pTau181 (B) in pg/mL are represented in the conversion to AD (-) and (+) populations. (C) ROC curves of plasma pTau217, pTau181 and CSF pTau181 and Abeta42/40 in MCI according to conversion to AD. Respective AUC (95% Confidence interval): plasma pTau217: 0.722 (0.657 to 0.781), plasma pTau181 0.673 (0.606 to 0.736), CSF pTau181: 0.712 (0.646 to 0.771), and CSF Aβ42/40: 0.733 (0.668 to 0.791).
